# Pandemic inequity in a megacity: a multilevel analysis of individual, community and health care vulnerability risks for COVID-19 mortality in Jakarta, Indonesia

**DOI:** 10.1101/2021.11.24.21266809

**Authors:** Henry Surendra, Ngabila Salama, Karina D Lestari, Verry Adrian, Widyastuti, Dwi Oktavia, Rosa N Lina, Bimandra A Djaafara, Ihsan Fadilah, Rahmat Sagara, Lenny L Ekawati, Ahmad Nurhasim, Riris A Ahmad, Aria Kekalih, Ari F Syam, Anuraj H Shankar, Guy Thwaites, J. Kevin Baird, Raph L. Hamers, Iqbal RF Elyazar

## Abstract

**Background:** The 33 recognized megacities comprise approximately 7% of the global population, yet account for 20% COVID-19 deaths. The specific inequities and other factors within megacities that affect vulnerability to COVID-19 mortality remain poorly defined. We assessed individual, community-level and health care factors associated with COVID-19-related mortality in a megacity of Jakarta, Indonesia, during two epidemic waves spanning March 2, 2020, to August 31, 2021.

**Methods:** This retrospective cohort included all residents of Jakarta, Indonesia, with PCR-confirmed COVID-19. We extracted demographic, clinical, outcome (recovered or died), vaccine coverage data, and disease prevalence from Jakarta Health Office surveillance records, and collected sub-district level socio-demographics data from various official sources. We used multi-level logistic regression to examine individual, community and sub-district-level health care factors and their associations with COVID-19-mortality.

**Findings:** Of 705,503 cases with a definitive outcome by August 31, 2021, 694,706 (98·5%) recovered and 10,797 (1·5%) died. The median age was 36 years (IQR 24–50), 13·2% (93,459) were <18 years, and 51·6% were female. The sub-district level accounted for 1·5% of variance in mortality (p<0.0001). Individual-level factors associated with death were older age, male sex, comorbidities, and, during the first wave, age <5 years (adjusted odds ratio (aOR) 1·56, 95%CI 1·04-2·35; reference: age 20-29 years). Community-level factors associated with death were poverty (aOR for the poorer quarter 1·35, 95%CI 1·17-1·55; reference: wealthiest quarter), high population density (aOR for the highest density 1·34, 95%CI 1·14-2·58; reference: the lowest), low vaccine coverage (aOR for the lowest coverage 1·25, 95%CI 1·13-1·38; reference: the highest).

**Interpretation:** In addition to individual risk factors, living in areas with high poverty and density, and low health care performance further increase the vulnerability of communities to COVID-19-associated death in urban low-resource settings.

**Funding:** Wellcome (UK) Africa Asia Programme Vietnam (106680/Z/14/Z).

**Research in context:** *Evidence before this study:* We searched PubMed on November 22, 2021, for articles that assessed individual, community, and healthcare vulnerability factors associated with coronavirus disease 2019 (COVID-19) mortality, using the search terms (“novel coronavirus” OR “SARS-CoV-2” OR “COVID-19”) AND (“death” OR “mortality” OR “deceased”) AND (“community” OR “social”) AND (“healthcare” OR “health system”). The 33 recognized megacities comprise approximately 7% of the global population, yet account for 20% COVID-19 deaths. The specific inequities and other factors within megacities that affect vulnerability to COVID-19 mortality remain poorly defined. At individual-level, studies have shown COVID-19-related mortality to be associated with older age and common underlying chronic co-morbidities including hypertension, diabetes, obesity, cardiac disease, chronic kidney disease and liver disease. Only few studies from North America, and South America have reported the association between lower community-level socio-economic status and healthcare performance with increased risk of COVID-19-related death. We found no studies have been done to assess individual, community, and healthcare vulnerability factors associated with COVID-19 mortality risk, especially in lower-and middle-income countries (LMIC) where accessing quality health care services is often challenging for substantial proportions of population, due to under-resourced and fragile health systems. In Southeast Asia, by November 22, 2021, COVID-19 case fatality rate had been reported at 2·2% (23,951/1,104,835) in Vietnam, 1·7% (47,288/2,826,853) in Philippines, 1·0% (20,434/2,071,009) in Thailand, 1·2% (30,063/2,591,486) in Malaysia, 2·4% (2,905/119,904) in Cambodia, and 0·3% in Singapore (667/253,649). Indonesia has the highest number of COVID-19 cases and deaths in the region, reporting 3·4% case fatality rate (143,744 /4,253,598), with the highest number of cases in the capital city of Jakarta. A preliminary analysis of the first five months of surveillance in Jakarta found that 497 of 4265 (12%) hospitalised patients had died, associated with older age, male sex; pre-existing hypertension, diabetes, or chronic kidney disease; clinical diagnosis of pneumonia; multiple (>3) symptoms; immediate intensive care unit admission, or intubation.

*Added value of this study:* This retrospective population-based study of the complete epidemiological surveillance data of Jakarta during the first eighteen months of the epidemic is the largest studies in LMIC to date, that comprehensively analysed the individual, community, and healthcare vulnerability associated with COVID-19-related mortality among individuals diagnosed with PCR-confirmed COVID-19. The overall case fatality rate among general population in Jakarta was 1·5% (10,797/705,503). Individual factors associated with risk of death were older age, male sex, comorbidities, and, during the first wave, age <5 years (adjusted odds ratio (aOR) 1·56, 95%CI 1·04-2·35; reference: age 20-29 years). The risk of death was further increased for people living in sub-districts with high rates of poverty (aOR for the poorer quarter 1·35, 95%CI 1·17-1·55; reference: wealthiest quarter), high population density (aOR for the highest density 1·34, 95%CI 1·14-2·58), and low COVID-19 vaccination coverage (aOR for the lowest coverage 1·25, 95%CI 1·13-1·38; reference: the highest).

*Implications of all available evidence:* Differences in socio-demographics and access to quality health services, among other factors, greatly influence COVID-19 mortality in low-resource settings. This study affirmed that in addition to well-known individual risk factors, community-level socio-demographics and healthcare factors further increase the vulnerability of communities to die from COVID-19 in urban low-resource settings. These results highlight the need for accelerated vaccine rollout and additional preventive interventions to protect the urban poor who are most vulnerable to dying from COVID-19.

## Background

There are currently 33 megacities, defined by the United Nations Department of Economic and Social Affairs as cities with a population of at least 10 million persons^1^. Megacities comprise 8% of the global population, yet account for approximately 20% of all COVID-19 deaths^2^. Megacities often contain high levels of inequity with regard to income, housing, sanitation, transportation, population density, basic health care, and other factors. The important role of health inequity in the spread and mortality of epidemics has been known from influenza in 1918 to Ebola in 2014^3–7^. The severity of illness and clinical outcomes can be affected by the concentration of comorbidities in susceptible groups in communities^3,4,8^, and through disparities of access to health care for preventive measures or prompt diagnosis and treatment. Ensuring health equity, especially in megacities experiencing massive urbanization and mobility is essential for the current and future global health threats.

In the context of the ongoing pandemic, understanding community-level risk factors associated with the mortality is very important to guide policymaking and target public health and clinical interventions, particularly in the context of fragile public health systems. At individual-level, older age and pre-existing chronic comorbidities have been consistently reported as the main risk factors of COVID-19-related mortality across different settings^9–13^. At the community-level, recent findings in US, Chile and Brazil suggested that COVID-19 mortality was concentrated in groups with higher socio-demographics vulnerability^14–18^. However, there is a general scarcity of data in LMIC assessing the influence of community-level socio-demographics factors on COVID-19-related mortality.

Indonesia, the world’s fourth most populous country (population 274 million), is a lower-middle income country (LMIC) featuring great geographic, cultural and socio-economic diversity across the archipelago. For example, the 2019 human development index (HDI) ranged from 0·32 in Nduga District, Papua to 0·87 in Yogyakarta city, Yogyakarta^19^. Indonesia has suffered the highest number of COVID-19 confirmed cases and deaths in Southeast Asia, second only to India in all of Asia^20^, at 4,253,598 cases and 143,744 deaths (3·4% case fatality rate (CFR)) up to November 22, 2021^21^, of which 20% (863,482) of cases and 9·4% (13,574) of deaths occurred in its capital Jakarta, a megacity (7,659 Km^2^, and estimated population 10·6 million) that features stark health inequalities and socio-demographic heterogeneity. The first SARS-CoV-2 epidemic wave occurred from March 2 2020 to April 30 2021, and a more intense second wave dominated by Delta variant peaked in July 2021^22,23^.

As in many LMIC, accessing quality health care services is challenging to substantial proportions of the Indonesian population, due to under-resourced and fragile health systems^24^. The 2018 public health development index (PHDI) ranges from 35% in Paniai district, Papua province to 75% in Gianyar district, Bali province. Within the province-level administration area called the Special Capital City Area Jakarta (*Daerah Khusus Ibukota*, DKI Jakarta), the PHDI ranged from 64% in North Jakarta to 68% in East Jakarta districts. However, the five districts of DKI Jakarta (North, East, West, South and Central) are highly heterogeneous socio-demographically and little is known regarding the capacity and performance of public health systems at sub-district level. That heterogeneity and the large number of COVID-19 cases and deaths during the first and second wave of the epidemic in DKI Jakarta provides insights directly relevant to the national public health response to the COVID-19 crisis, as well as other LMIC settings. In this study, we assessed individual, community-level and health care vulnerability among the 44 sub-districts of DKI Jakarta and how those factors were associated with COVID-19-related mortality during the first 18 months of the epidemic in that province (March 2020 through August 2021).

## Methods

### Study design and participants

This was a retrospective cohort study of all adults and children diagnosed with PCR-confirmed SARS-CoV-2 infection (COVID-19 cases) in DKI Jakarta, Indonesia, recorded by the DKI Jakarta Health Office, who either died or recovered between March 2, 2020 and August 31, 2021. We restricted the analysis to DKI Jakarta residents to enable robust estimations of community-level risk factors and individual outcomes (deceased vs recovered) of cases living in the corresponding sub-district (Figure 1A). In accordance with Indonesia’s national COVID-19 guidelines^25^, confirmatory SARS-CoV-2 PCR testing was conducted on naso-and/or oropharyngeal swab specimens in COVID-19 reference laboratories.

**Fig. 1.**
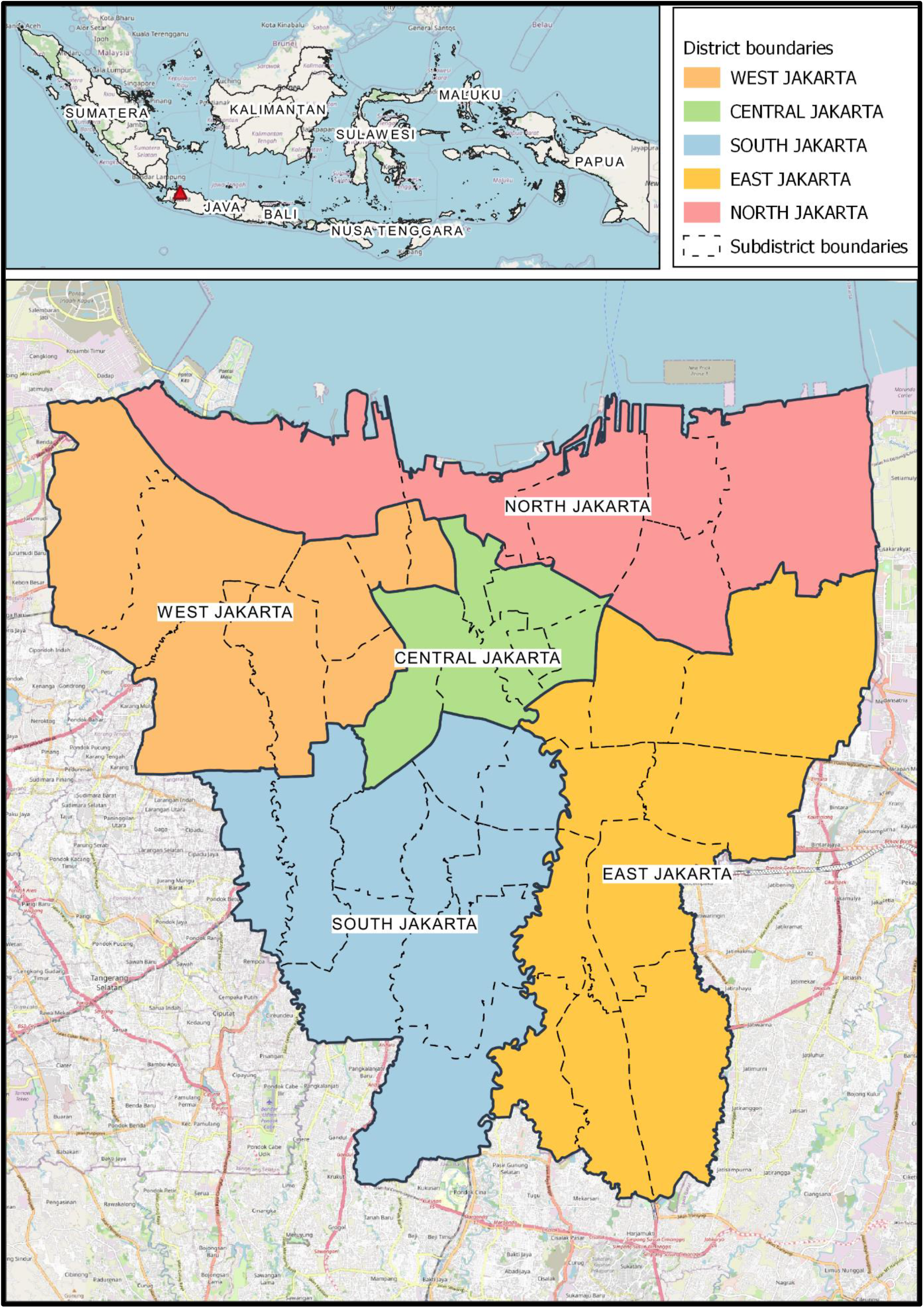
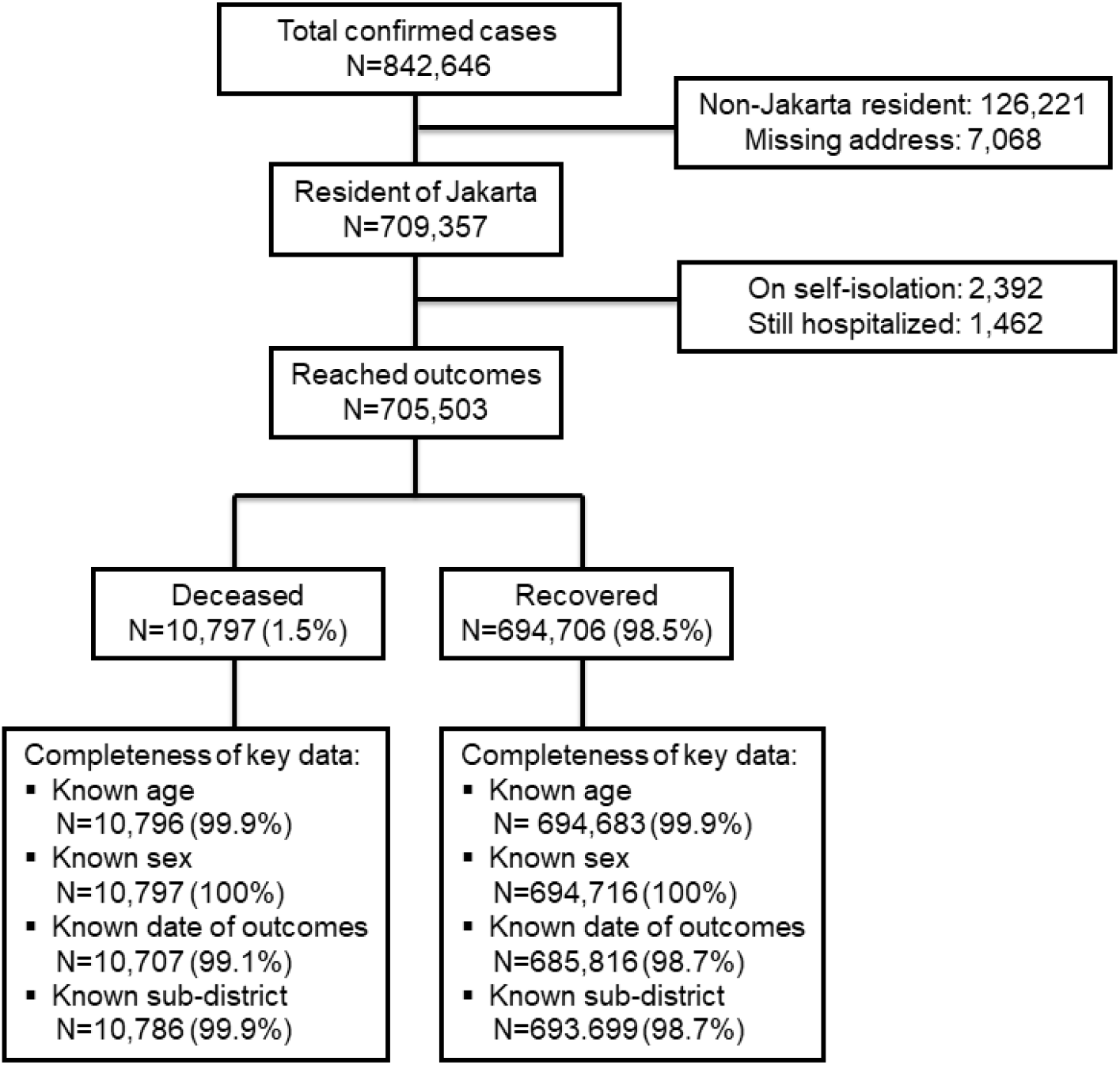
Study sites (A), and flow chart and completeness of key variables (B).

### Data collection and operational definitions

Individual-level data were collected from all cases who lived in any of 44 sub-districts in DKI Jakarta. Each sub-district public health facility had designated epidemiologists and surveillance officers responsible for epidemiological investigations using an official COVID-19 case investigation form capturing demographical and clinical data of each confirmed case. Completed forms were submitted to the DKI Jakarta Health Office for cleaning and verification (checking for completeness, inconsistency, error, and duplication) and entered into a surveillance database. We extracted data regarding SARS-CoV-2 PCR testing, hospital admission, and outcomes (recovered or deceased), along with age, sex, and pre-existing comorbidities (based on clinical assessment or cases self-report)^25^.

Sub-district-level data (community-level risk factors) were collected from official government websites. Data on number of population, population density (number of residents per square kilometre (Km^2^), number of neighbourhoods with poor sanitation, number of nurses, and number of medical doctors were collected from the DKI Jakarta Government Integrated Data Portal^26^. Data on the number of individuals categorised as poorest (the lowest tenth of the national level of poverty) were obtained from the National Team for the Acceleration of Poverty Reduction, and used to calculate proportion of poorest individuals by sub-district^27^. Population density, poverty, and proportion of poor sanitation areas were used to describe socio-demographic vulnerability. Data on COVID-19 vaccine coverage, universal child immunization coverage, all-cause mortality among under 5 years population, prevalence of hypertension, diabetes, and tuberculosis were collected from the DKI Jakarta Health Office surveillance records. Doctor-population ratio (number of doctors per 10,000 population in 2020), nurse-population ratio (number of nurses per 10,000 population in 2020), COVID-19 vaccine coverage (proportion of individuals received two-dose of COVID-19 vaccine per August 31 2021), universal child immunization coverage (proportion of children received complete dosage of government mandatory vaccination), all-cause mortality among under 5 years population (proportion of all of deaths per 1000 under 5 years population), prevalence of hypertension in 2019 (proportion of cases per 100 population), prevalence of diabetes in 2019 (proportion of cases per 100 population), and prevalence of tuberculosis in 2019 (proportion of cases per 100 population) by sub-district were used to describe health care vulnerability.

### Statistical analysis

Numeric values of each community-level risk factors were categorised into quarters, i.e., below 25^th^ percentile (quarter 1), 25-50^th^ percentile (quarter 2), 50-75^th^ percentile (quarter 3) and above 75^th^ percentile (quarter 4). Descriptive statistics included proportions for categorical variables and medians and interquartile ranges (IQRs) for continuous variables. We used the Mann-Whitney U test, χ^2^ test, or Fisher’s exact test to compare characteristics between deceased and recovered cases. We set statistical significance at 0·05, and all tests were two sided.

We used bivariable and multivariable multi-level logistic regression models to determine the risk of death, expressed as odds ratio (OR) with 95% confidential intervals (CI). Sub-district was treated as the random effect variable to adjust for clustering of observations within sub-districts. We did null model analysis (no predictor was added) and the result justified the use of the multi-level models. We excluded cases from two sub-districts with insufficient sample size (Kepulauan Seribu Selatan and Utara). All independent variables with p-value <0·10 in bivariable analysis were included in the multivariable models. Final model selection was informed by intra class correlation postestimation test. We used interaction terms to examine potential effect modification by age, sex, and time. In the presence of interaction, the stratum-specific OR and 95% CI were calculated, adjusting for other variables with p-value <0·10 in bivariable analysis. Additionally, we used a restricted cubic spline mixed effect regression to model the OR of death over time.

There was a substantial proportion of missing data for chronic comorbidities (58%). Missing-indicator analysis by risk factor stratification and by regression analysis identified bias of missing data with respect to mortality, thus we additionally conducted analysis to assess sensitivity of risk factor identification due to missing data. We performed multi-level logistic regression analysis with multiple imputations (100 imputed datasets), treating sub-district as random effect variable. Age, sex, and outcome were included as independent variables in imputing the comorbidities variable. All statistical analyses were done in Stata/MP 17.1 (StataCorp, College Station, TX, USA). This study is reported as per Strengthening the Reporting of Observational Studies in Epidemiology (STROBE) guidelines.

### Ethics

This study was approved by the Health Research Ethics Committee of the National Institute of Health Research and Development, Ministry of Health Indonesia (LB.02.01/2/KE.643/2021). The requirement for patient consent was waived as this was a secondary analysis of anonymised routine surveillance data^21^.

## Results

Between March 2 and August 31, 2021, a total of 842,646 PCR-confirmed COVID-19 cases were recorded by the DKI Jakarta Health Office (Figure 1). Of those, 709,357 (84·2%) lived in DKI Jakarta and 705,503 (99·5%) had reached a definitive outcome before September 1, 2021, i.e., those deceased or recovered and were included in this analysis. The 2392 (0·3%) individuals who were still hospitalised, and 1462 (0·2%) who were in self-isolation were not included in analysis. The study flow chart and completeness of key data are presented in Figure 1B.

Table 1 presents the characteristics of the 705,503 cases included in the analysis. The median age was 36 years (IQR 24-50, range 0·1-121), with 93,459 (13·2%) under 18 years, and 364,133 (51·6%) were women, and 233,025 (33·0%) had been hospitalised due to COVD-19. The second wave of the pandemic comprised 372,688 (53·7%) cases, with 288,228 (40·9%) having no chronic comorbidities, 4,974 (0·7%) with at least one comorbidity, and 412,301 (58·4%) with unknown status of comorbidities.

**Table 1:** Individual, community, health care characteristics and outcomes of COVID-19 cases in DKI Jakarta, March 2, 2020 to August 31, 2021.

|  | Total<br>n=705,503 | Deceased<br>n=10,797 | Recovered<br>n=694,706 | p value |
| --- | --- | --- | --- | --- |
| **Individual-level characteristics** |  |  |  |  |
| Median age (IQR), years | 36 (24–50) | 59 (50-68) | 35 (24-49) | <0.0001 |
| Age group, years |  |  |  | <0.0001 |
| 0-4 | 21,793 (3.1%) | 47 (0.4%) | 21,746 (3.1%) |  |
| 5-9 | 23,070 (3.3%) | 16 (0.2%) | 23,054 (3.3%) |  |
| 10-19 | 66,514 (9.4%) | 72 (0.7%) | 66,514 (9.4%) |  |
| 20-29 | 149,267 (21.2%) | 311 (2.9%) | 148,956 (21.4%) |  |
| 30-39 | 146,900 (20.8%) | 692 (6.4%) | 146,208 (21.1%) |  |
| 40-49 | 121,454 (17.2%) | 1,455 (13.5%) | 119,999 (17.2%) |  |
| 50-59 | 98,934 (14.0%) | 3,115 (28.9%) | 95,819 (13.8%) |  |
| 60-69 | 52,776 (7.5%) | 2,827 (26.2%) | 49,949 (7.2%) |  |
| ≥70 | 24,761 (3.5%) | 2,261 (20.9%) | 22,500 (3.2%) |  |
| Sex |  |  |  | <0.0001 |
| Female | 364,133 (51.6%) | 4,810 (44.6%) | 359,323 (51.7%) |  |
| Male | 341,370 (48.4%) | 5,987 (55.4%) | 335,383 (48.3%) |  |
| Hospitalised |  |  |  | <0.0001 |
| No | 472,478 (67.0%) | 105 (1.0%) | 427,373 (68.0%) |  |
| Yes | 233,025 (33.0%) | 10,692 (99.0%) | 222,333 (32.0%) |  |
| Comorbidities |  |  |  | <0.0001 |
| Absent | 288,228 (40.9%) | 2,754 (25.5%) | 285,474 (41.1%) |  |
| Present | 4,974 (0.7%) | 468 (4.3%) | 4,506 (0.7%) |  |
| Unknown | 412,301 (58.4%) | 7,575 (70.2%) | 404,726 (58.3%) |  |
| Period of time |  |  |  | <0.0001 |
| First wave | 321,734 (46.3%) | 6,107 (57.1%) | 315,627 (46.2%) |  |
| Second wave | 372,688 (53.7%) | 4,585 (42.9%) | 368,103 (53.8%) |  |
| **Sub-district-level characteristics\*** |  |  |  |  |
| **Socio-demographics** |  |  |  |  |
| Population density,<br>population/Km<sup>2</sup> |  |  |  | <0.0001 |
| Q1 (<14,093) | 157,561 (22.4%) | 2,164 (20.1%) | 155,397 (22.4%) |  |
| Q2 (14,093-17,092) | 199,848 (28.4%) | 2,939 (27.3%) | 196,909 (28.4%) |  |
| Q3 (17,092-22,578) | 194,519 (27.6%) | 3,154 (29.2%) | 191,365 (27.6%) |  |
| Q4 (22,578-50,829) | 152,557 (21.6%) | 2,529 (23.4%) | 150,028 (21.6%) |  |
| Poverty, % |  |  |  | <0.0001 |
| Q1 (0.2-1.5) | 199,916 (28.4%) | 2,717 (25.2%) | 197,199 (28.4%) |  |
| Q2 (1.5-2.1) | 156,153 (22.1%) | 2,614 (24.2%) | 153,539 (22.1%) |  |
| Q3 (2.1-3.6) | 173,167 (24.6%) | 2,712 (25.2%) | 170,455 (24.6%) |  |
| Q4 (3.6-8.8) | 175,249 (24.9%) | 2,743 (25.4%) | 172,506 (24.9%) |  |
| Poor sanitation areas, % |  |  |  | 0.135 |
| Q1 (0-47.9) | 181,868 (25.8%) | 2,747 (25.5%) | 179,121 (25.8%) |  |
| Q2 (47.9-59.9) | 191,093 (27.2%) | 2,846 (26.4%) | 188,247 (27.2%) |  |
| Q3 (59.9-63.8) | 158,002 (22.4%) | 2,464 (22.8%) | 155,538 (22.4%) |  |
| Q4 (63.8-89.2) | 173,522 (24.6%) | 2,729 (25.3%) | 170,793 (24.6%) |  |
| **Health care capacity** |  |  |  |  |
| Doctor-population ratio, doctor per<br>10,000 population |  |  |  | 0.158 |
| Q1 (2.6-5.2) | 177,254 (25.2%) | 2,680 (24.8%) | 174,574 (25.2%) |  |
| Q2 (5.2-7.9) | 191,918 (27.2%) | 3,012 (27.9%) | 188,906 (27.2%) |  |
| Q3 (7.9-9.9) | 166,147 (23.6%) | 2,467 (22.9%) | 163,680 (23.6%) |  |
| Q4 (9.9-74.8) | 169,166 (24.0%) | 2,627 (24.4%) | 166,539 (24.0%) |  |
| Nurse-population ratio, nurse per<br>10,000 population |  |  |  | 0.007 |
| Q1 (2.9-11.2) | 185,251 (26.3%) | 2,755 (25.5%) | 182,496 (26.3%) |  |
| Q2 (11.2-22.8) | 161,767 (23.0%) | 2,553 (23.7%) | 159,214 (23.0%) |  |
| Q3 (22.8-83.9) | 176,863 (25.1%) | 2,611 (24.2%) | 174,252 (25.1%) |  |
| Q4 (83.9-416.1) | 180,604 (25.6%) | 2,867 (26.6%) | 177,737 (25.6%) |  |
| COVID-19 vaccination coverage, % |  |  |  | <0.0001 |
| Q1 (33.0-36.1) | 190,502 (27.0%) | 3,254 (30.2%) | 187,248 (27.0%) |  |
| Q2 (36.1-38.5) | 164,821 (23.4%) | 2,662 (24.7%) | 162,159 (23.4%) |  |
| Q3 (38.5-40.5) | 172,210 (24.5%) | 2,358 (21.8%) | 169,852 (24.5%) |  |
| Q4 (40.5-50.0) | 176,952 (25.1%) | 2,512 (23.3%) | 174,440 (25.1%) |  |
| Universal child immunization coverage, % |  |  |  | <0.0001 |
| Q1 (94.5-96.2) | 180,820 (25.7%) | 2,682 (24.9%) | 178,138 (25.7%) |  |
| Q2 (96.2-98.0) | 191,987 (27.2%) | 2,862 (26.5%) | 189,125 (27.2%) |  |
| Q3 (98.0-98.9) | 155,023 (22.0%) | 2,586 (24.0%) | 152,437 (22.0%) |  |
| Q4 (98.9-100.0) | 173,999 (25.1%) | 2,656 (24.6%) | 173,999 (25.1%) |  |
| **Health related characteristics** |  |  |  |  |
| Prevalence of Hypertension, % |  |  |  | 0.490 |
| Q1 (4.2-7.1) | 173,268 (24.6%) | 2,693 (25.0%) | 170,575 (24.6%) |  |
| Q2 (7.1-8.7) | 203,932 (29.0%) | 3,127 (29.0%) | 200,805 (28.9%) |  |
| Q3 (8.7-13.6) | 145,395 (20.6%) | 2,165 (20.0%) | 143,230 (20.7%) |  |
| Q4 (13.6-33.0) | 181,890 (25.8%) | 2,801 (26.0%) | 179,089 (25.8%) |  |
| Prevalence of Diabetes, % |  |  |  | <0.0001 |
| Q1 (0.6-1.3) | 188,051 (26.0%) | 2,934 (27.2%) | 180,117 (26.0%) |  |
| Q2 (1.3-2.1) | 160,975 (22.9%) | 2,284 (21.2%) | 158,691 (22.9%) |  |
| Q3 (2.1-3.2) | 176,348 (25.0%) | 2,682 (24.9%) | 173,666 (25.0%) |  |
| Q4 (3.2-5.4) | 184,111 (26.1%) | 2,886 (26.7%) | 181,225 (26.1%) |  |
| Prevalence of Tuberculosis, % |  |  |  | <0.0001 |
| Q1 (0.1-0.2) | 181,457 (25.8%) | 2,504 (23.2%) | 178,953 (25.8%) |  |
| Q2 (0.2-0.3) | 188,309 (26.7%) | 2,865 (26.6%) | 185,444 (26.7%) |  |
| Q3 (0.3-0.5) | 171,319 (24.3%) | 2,741 (25.4%) | 168,578 (24.3%) |  |
| Q4 (0.5-2.4) | 163,400 (23.2%) | 2,676 (24.8%) | 160,724 (23.2%) |  |
| All-cause mortality per 1000 under 5 years old population |  |  |  | <0.0001 |
| Q1 (0.8-2.1) | 201,666 (28.6%) | 3,167 (29.4%) | 198,499 (28.6%) |  |
| Q2 (2.1-2.6) | 137,785 (19.6%) | 2,054 (19.0%) | 135,731 (19.6%) |  |
| Q3 (2.6-4.0) | 204,265 (29.0%) | 2,872 (26.6%) | 201,393 (29.0%) |  |
| Q4 (4.0-6.3) | 160,769 (22.8%) | 2,693 (25.0%) | 158,076 (22.8%) |  |

Regarding socio-demographics, 150,028 (21·6%) cases lived in sub-districts with the highest population density (22,578-50,829 people/Km^2^), and 175,249 (24·9%) with the highest proportion of poor population (3·6-8·8%). Regarding health care, 190,502 (27·0%) lived in the lowest COVID-19 vaccine coverage (33·0-36·1%), 181,890 (25·8%) lived in the highest hypertension prevalence (13·6-33·0%), 184,111 (26·1%) in the highest diabetes prevalence (3·2-5·4%), 163,400 (23·2%) in the highest tuberculosis prevalence (0·5-2·4%), and 173,999 (25·1%) in the highest childhood vaccine coverage (98·9-100·0%) (See Table 1 for details).

Of the 705,503 cases with a known outcome, 694,706 (98·5%) had recovered, 10,797 (1·5%) had died, and 105 (1·0%) had been declared dead at home and without hospitalisation. The highest numbers of cases (39% of 705,503), and deaths (25% of 10,797) were observed in July 2021 (Supplementary Figure 1A). Although a large majority of deaths (76%, 8203) was 50 years or older, death occurred across all age groups. Age-specific CFRs were 0·2% (47/21,793) for <5 years; 0·1% (16/23,070) for 5-9 years; 0·1% (72/66,514) for 10-19 years; 0·2% (311/149,267) for 20-29 years; 0·5% (692/146,900) for 30-39 years; 1·2% (1455/121,454) for 40-49 years; 3·2% (3115/98,934) for 50-59 years; 5·4% (2827/52,776) for 60-69 years; and 9·1% (2261/24,761) for ≥70 years (Supplementary Table 1 and Supplementary Figure 1B).

Compared to recovered cases, deceased cases were older (median 59 vs 35 years); more likely to be males (1·8% vs 1·3%), to have one or more comorbidities (9·4% vs 1·0%), to be infected in the first wave (1·9% vs 1·2%), and to live in sub-districts with higher population density (highest density: 1·7% vs lowest density: 1·4%), higher poverty (highest: 1·6% vs lowest: 1·4%); higher nurse-population ratio (lowest: 1·6% vs highest: 1·5%); and lower vaccine coverage (lowest: 1·7% vs highest: 1·4%) (Table 1). Compared with the first wave, there was a notable decrease in CFRs across sub-districts during the second wave of the epidemic (Figure 2E-F, and Figure 3B). Moreover, the sub-districts with higher population density, poverty, and lower vaccine coverage tended to be the sub-districts with higher CFR (Figure 2A-D, and Figure 3C).

**Fig. 2.**
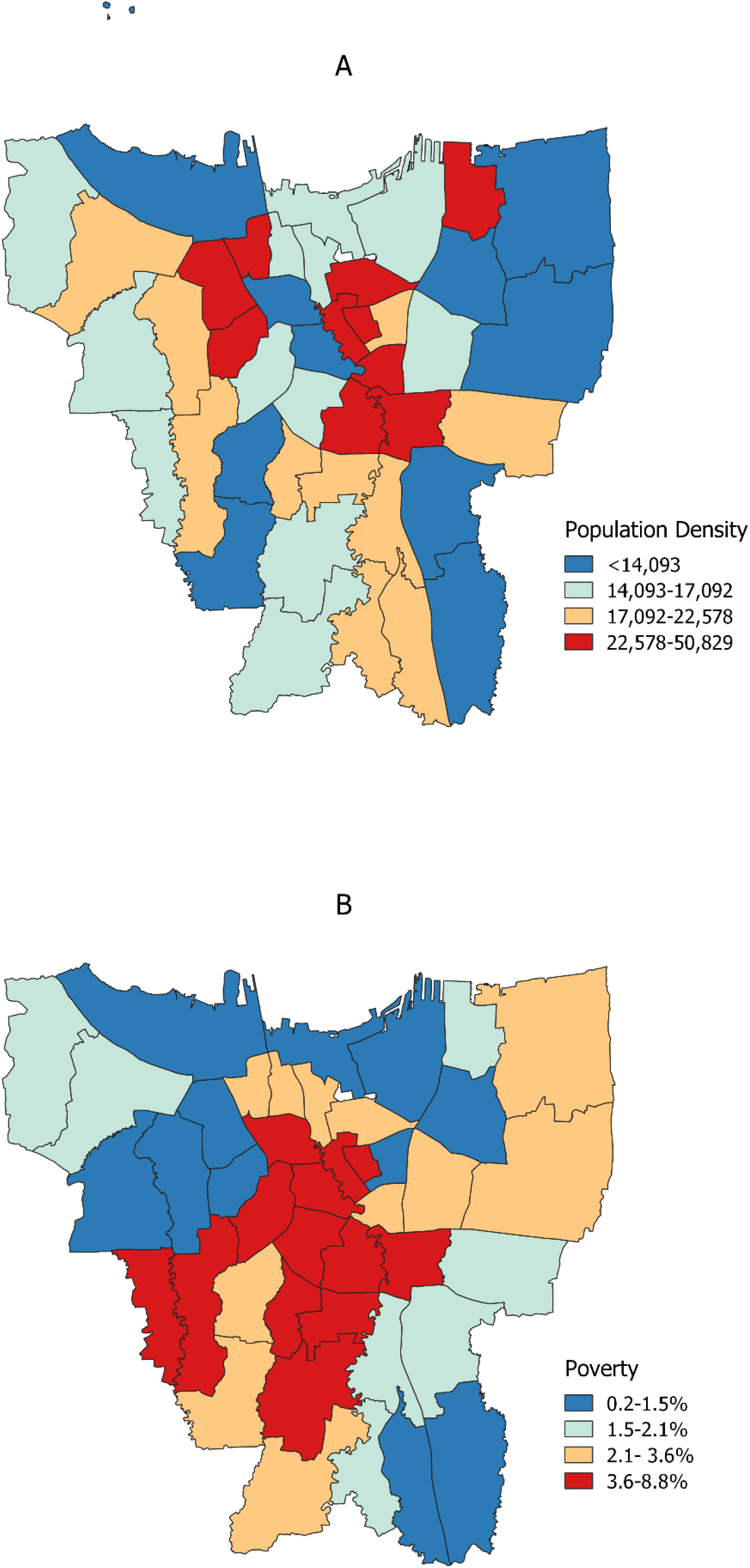
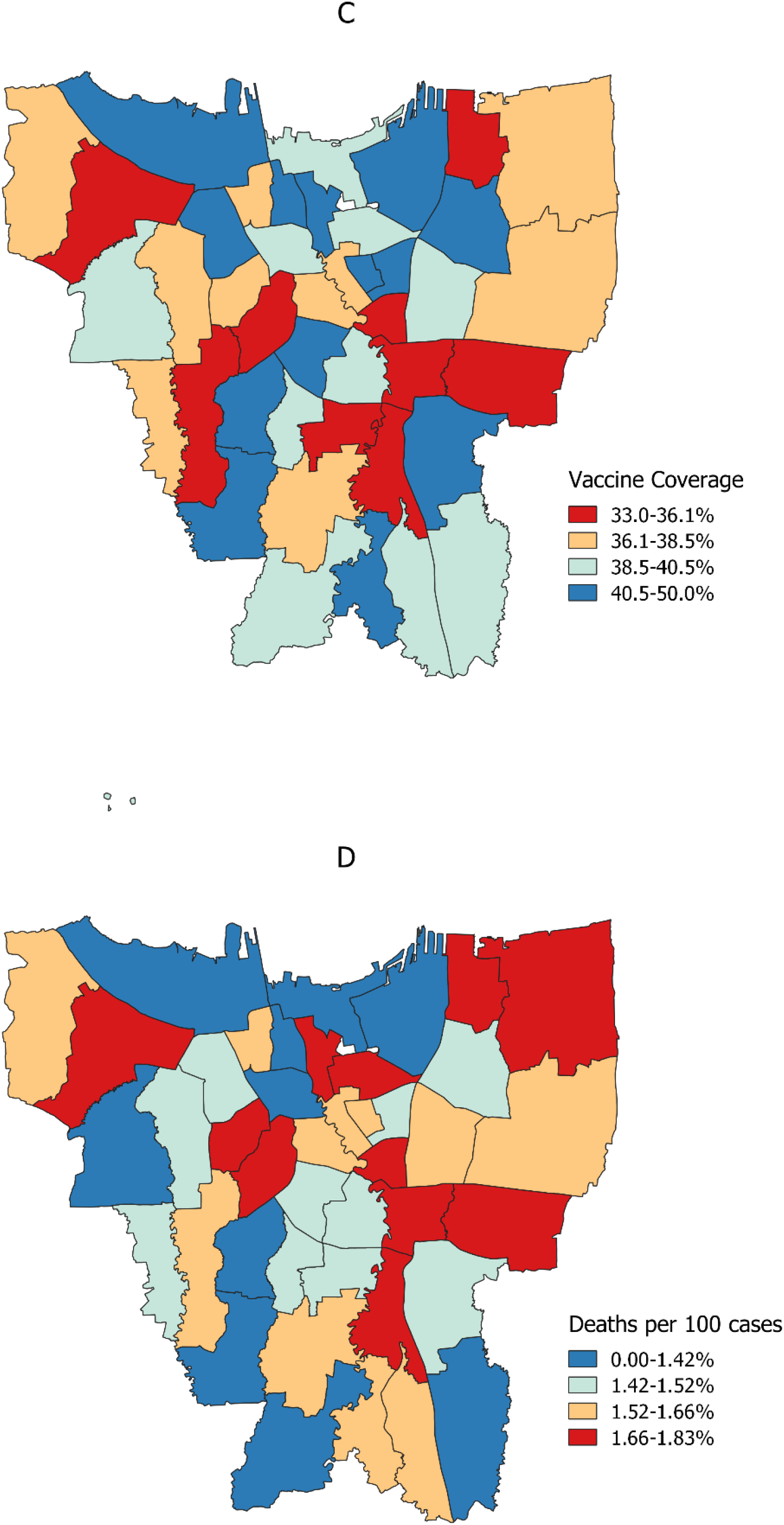
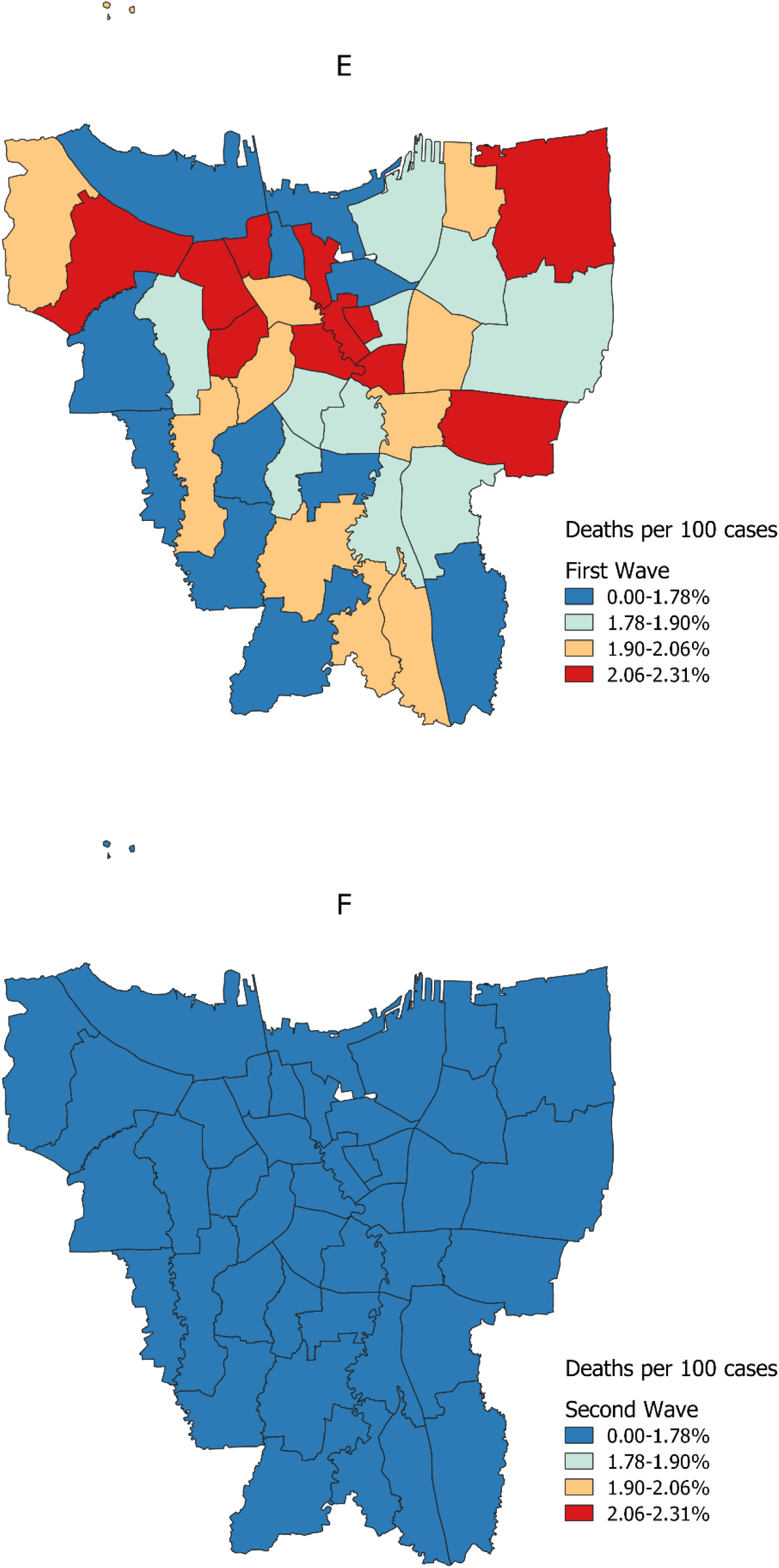
Characteristics of study sites. Sites categorised based on sub-district population density (A), poverty level (B), COVID-19 vaccine coverage per August 31 2021 (C), overall case fatality rate (D), case fatality rate during the first wave (E), and case fatality rate during the second wave (F). Black lines represent the sub-district administrative border.

**Fig. 3.**
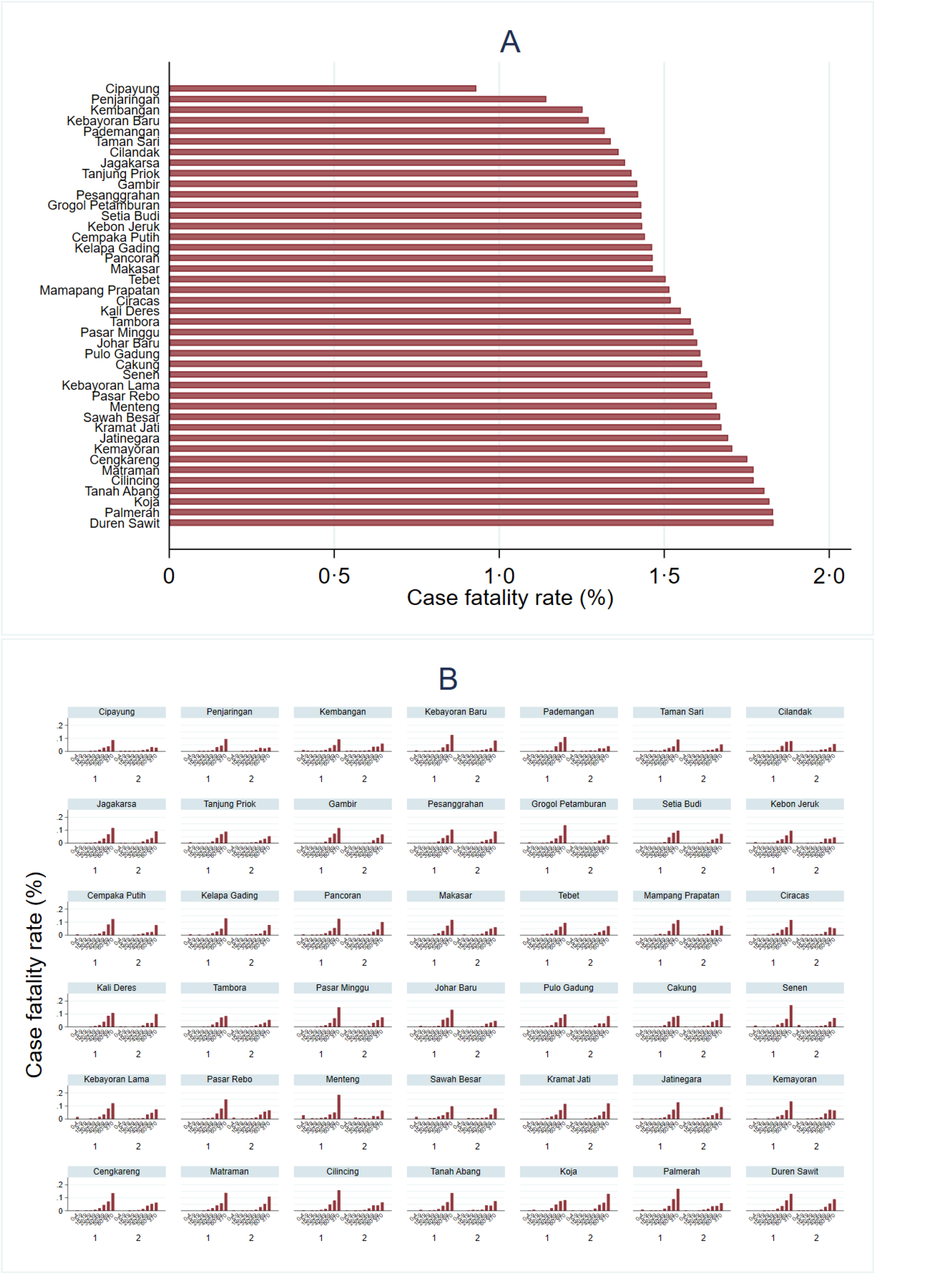
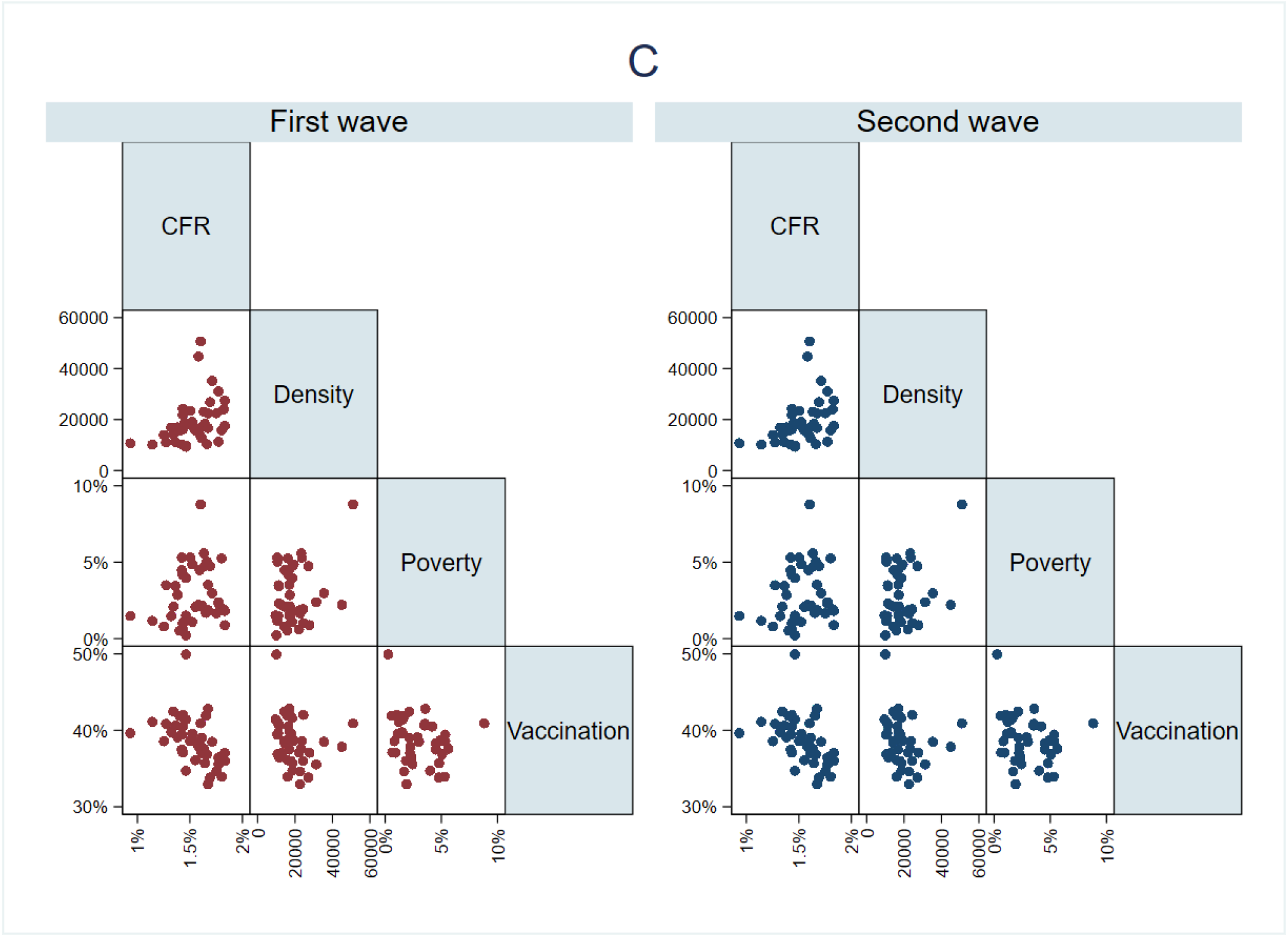
Overall case fatality rate (CFR) by sub-district (A), age-specific CFR per sub-district and by pandemic wave (B), and correlation between CFR with sub-district population density, poverty, and COVID-19 vaccine coverage by epidemic wave (C).

In bivariable analysis (Table 2), the risk of death was significantly associated with older age, male sex, comorbidities, first wave, and higher sub-district population density, poverty, higher prevalence of tuberculosis, all-cause mortality among under 5 years, and lower COVID-19 vaccine coverage.

**Table 2:** Bivariable analysis of individual, community, and health care risk factors associated with COVID-19 mortality in DKI Jakarta, March 2, 2020 to August 31, 2021.

|  | Crude OR (95% CI) | p value |
| --- | --- | --- |
| **Individual-level factors** |  |  |
| Age group, years |  |  |
| 0-4 | 1.03 (0.76-1.40) | 0.836 |
| 5-9 | 0.33 (0.20-0.55) | <0.0001 |
| 10-19 | 0.52 (0.40-0.67) | <0.0001 |
| 20-29 | 1 (ref) | .. |
| 30-39 | 2.27 (1.99-2.60) | <0.0001 |
| 40-49 | 5.81 (5.14-6.57) | <0.0001 |
| 50-59 | 15.59 (13.86-17.52) | <0.0001 |
| 60-69 | 27.26 (24.23-30.67) | <0.0001 |
| ≥70 | 49.18 (43.63-55.43) | <0.0001 |
| Sex |  |  |
| Female | 1 (ref) | .. |
| Male | 1.34 (1.28-1.39) | <0.0001 |
| Comorbidities |  |  |
| Present | 10.75 (9.70-11.91) | <0.0001 |
| Absent | 1 (ref) |  |
| Period of time |  |  |
| First wave | 1.55 (1.49-1.61) | <0.0001 |
| Second wave | 1 (ref) |  |
| **Socio-demographics** |  |  |
| Population density |  |  |
| Lowest | 1 (ref) | .. |
| Q2 | 1.06 (0.96-1.18) | 0.229 |
| Q3 | 1.16 (1.04-1.28) | 0.007 |
| Highest | 1.20 (1.08-1.33) | 0.001 |
| Poverty level |  |  |
| Lowest | 1 (ref) | .. |
| Q2 | 1.21 (1.09-1.34) | <0.0001 |
| Q3 | 1.15 (1.04-1.27) | 0.006 |
| Highest | 1.15 (1.04-1.27) | 0.005 |
| Proportion of poor sanitation areas |  |  |
| Lowest | 1 (ref) | .. |
| Q2 | 0.98 (0.87-1.10) | 0.744 |
| Q3 | 1.15 (1.04-1.27) | 0.526 |
| Highest | 1.15 (1.04-1.27) | 0.664 |
| **Health care capacity** |  |  |
| Vaccination coverage |  |  |
| Lowest | 1.20 (1.09-1.31) | <0.0001 |
| Q2 | 1.14 (1.04-1.24) | 0.005 |
| Q3 | 0.96 (0.88-1.06) | 0.423 |
| Highest | 1 (ref) | .. |
| Doctor-population ratio |  |  |
| Lowest | 0.97 (0.87-1.10) | 0.691 |
| Q2 | 1.01 (0.90-1.12) | 0.899 |
| Q3 | 0.95 (0.85-1.07) | 0.433 |
| Highest | 1 (ref) | .. |
| Nurse-population ratio |  |  |
| Lowest | 0.94 (0.84-1.05) | 0.292 |
| Q2 | 0.99 (0.88-1.11) | 0.831 |
| Q3 | 0.94 (0.84-1.05) | 0.278 |
| Highest | 1 (ref) | .. |
| Universal child immunization |  |  |
| Lowest | 1.00 (0.90-1.12) | 0.980 |
| Q2 | 1.10 (0.97-1.24) | 0.131 |
| Q3 | 1.03 (0.92-1.15) | 0.666 |
| Highest | 1 (ref) | .. |
| **Health related characteristics** |  |  |
| Prevalence of hypertension |  |  |
| Lowest | 1 (ref) | .. |
| Q2 | 0.97 (0.86-1.09) | 0.628 |
| Q3 | 0.97 (0.86-1.10) | 0.685 |
| Highest | 1.00 (0.89-1.12) | 0.999 |
| Prevalence of diabetes |  |  |
| Lowest | 1 (ref) | .. |
| Q2 | 0.89 (0.80-1.01) | 0.292 |
| Q3 | 0.96 (0.86-1.07) | 0.831 |
| Highest | 0.97 (0.86-1.08) | 0.278 |
| Prevalence of tuberculosis |  |  |
| Lowest | 1 (ref) | .. |
| Q2 | 1.10 (0.99-1.22) | 0.078 |
| Q3 | 1.12 (1.00-1.25) | 0.044 |
| Highest | 1.18 (1.06-1.31) | 0.003 |
| All-cause mortality among under 5 years old population |  |  |
| Lowest | 1 (ref) | .. |
| Q2 | 0.95 (0.86-1.06) | 0.398 |
| Q3 | 0.90 (0.82-1.00) | 0.045 |
| Highest | 1.07 (0.96-1.19) | 0.209 |

In the final multivariable multi-level logistic regression model (Figure 4A), the risk of death was increased for age groups 30-39 years (aOR 1·94, 95%CI 1·62-2·33), 40-49 years (aOR 4·51, 95%CI 3·82-5·33), 50-59 years (aOR 12·65, 95%CI 10·80-14·81), 60-69 years (aOR 18·64, 95%CI 15·87-21·89), ≥70 years (aOR 32·91, 95%CI 27·97-38·72) compared to 20-29 years; for males (aOR 1·29, 95%CI 1·24-1·34); for individuals with at least one comorbidity (aOR 3·96, 95%CI 3·56-4·41); for residents of sub-districts with highest population density (aOR 1·34, 95%CI 1·14-1·58, reference: lowest density), higher poverty (Q3) (aOR 1·35, 95%CI 1·17-1·55, reference: lowest poverty), and with lowest vaccine coverage (aOR 1·25, 95%CI 1·13-1·38, reference: highest coverage). We found no associations with proportion of poor sanitation areas, doctor-population ratio, nurse-population ratio, prevalence of hypertension, diabetes, and tuberculosis (p>0·05 each). The sensitivity analysis revealed similar findings, suggesting there was no significant bias introduced by missing data in our dataset; it also suggested that the risk of death was increased for cases who had at least one comorbidity (aOR 4·25, 95%CI 3·81-4·75) compared to those who had no comorbidity (Supplementary Table 2).

**Fig. 4.**
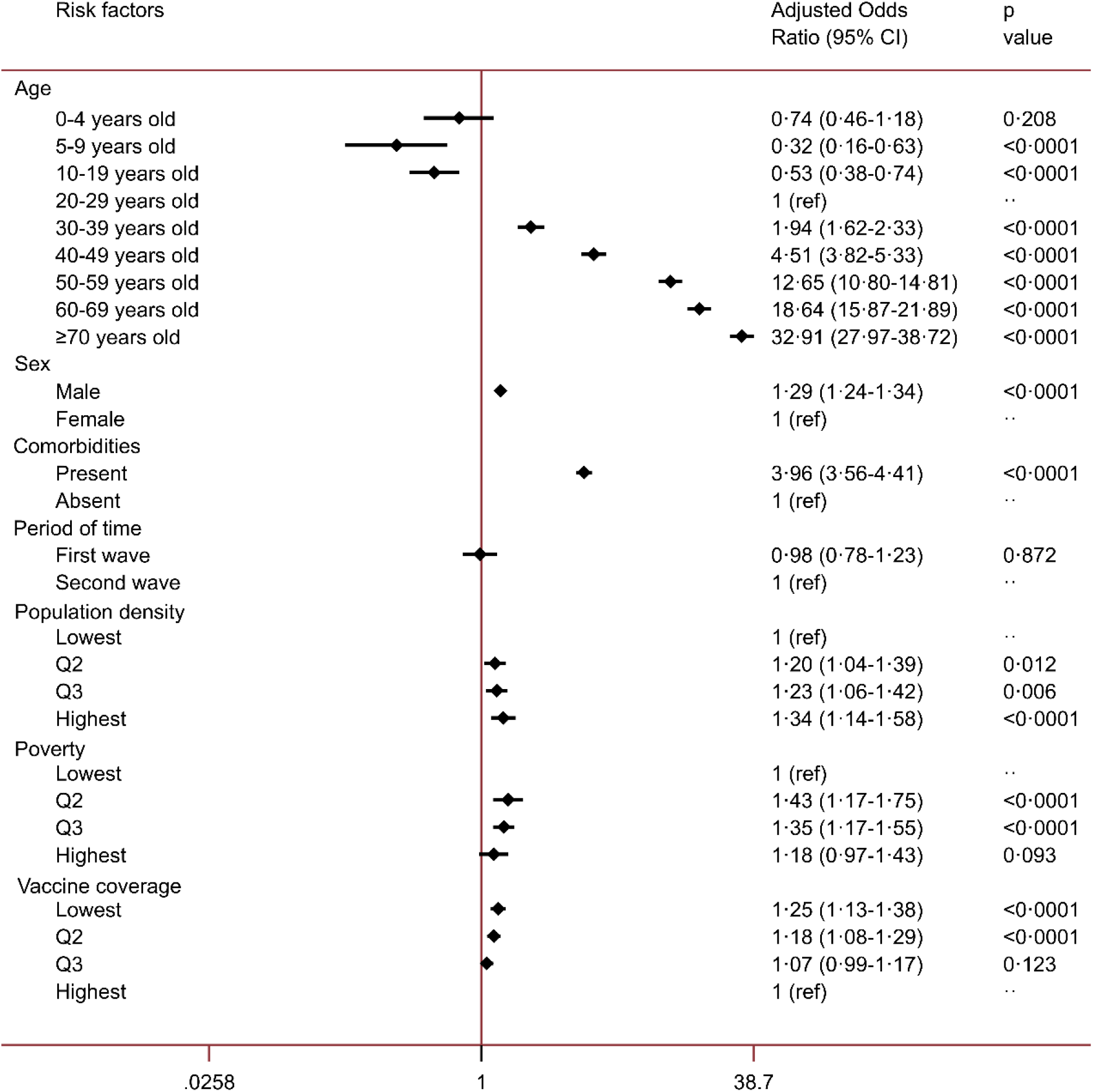
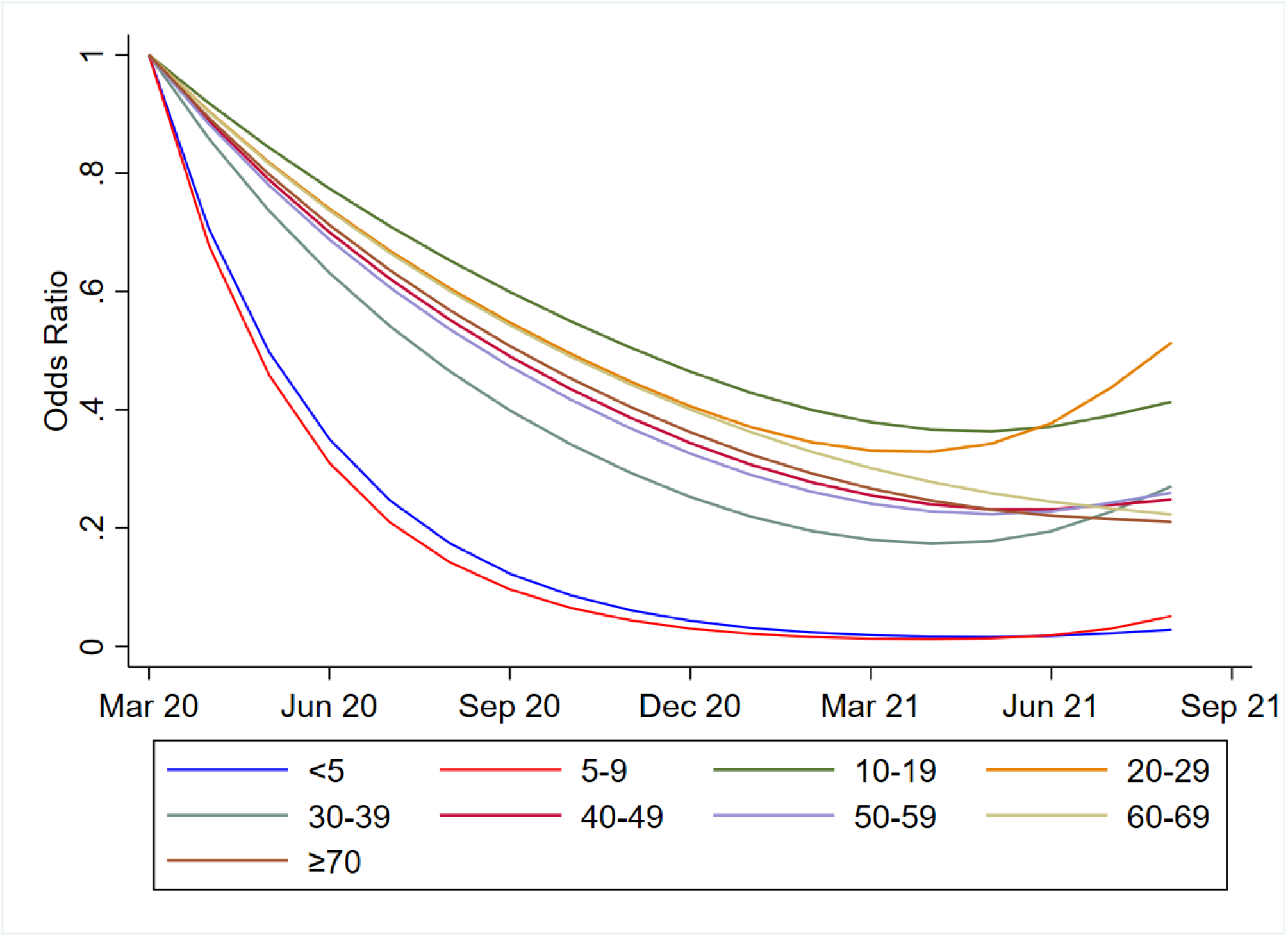
Final multi-level logistic regression model showing individual, community, and health care factors associated with COVID-19 mortality (A), and age-specific COVID-19 mortality risk over time (B) in DKI Jakarta, Indonesia, March 2, 2020 to August 31, 2021. Sub-district was treated as the random effect variable in both models. For analysis presented in Figure A, first wave=March 2020 to April 2021. Second wave=May 2021 to August 2021. Numeric values were categorised into quartiles (Q), i.e., below 25^th^ percentile (Lowest), 25^th^–50^th^ percentile (Q2), 50^th^–75^th^ percentile (Q3) and above 75^th^ percentile (Highest) for each sub-district level variable. For Figure B, each line represents age-specific odds ratio estimates obtained from restricted cubic spline multi-level logistic regression model.

We found that the effect of age was modified by time (first wave), and poverty was modified by population density (p<0·0001). Although higher age was associated with increased risk of death, we found that the risk of death was higher for children 0-4 years (aOR 1·56 95%CI 1·04-2·35) compared to adult age 20-29 years in the first pandemic wave, but not in the second wave (Supplementary Table 3). We found that mortality risk significantly decreased over time especially for children 0-9 years (Figure 4B). In addition, we found that the risk of death was higher for sub-districts with highest level of poverty and density compared to sub-districts with lowest poverty and density (aOR 1·27, 95%CI 1·10-1·47) (Supplementary Table 4). Sex was not found to be an effect modifier in the final model.

## Discussion

This retrospective study described the complete epidemiological surveillance data of PCR-confirmed COVID-19 cases in DKI Jakarta, including 705,503 adults and children living in 44 sub-districts, during two epidemic waves spanning the first 18 months of the SARS-CoV-2 transmission in Indonesia. This analysis represents the largest reported case series from any LMIC to date. The overall CFR was 1·2% (10,797/705,503), and deaths occurred across all ages. People aged less than 50 accounted for 75% of cases and 81% of the population, while those older than 50 accounted for 76% of deaths but only 19% of the population. Mortality increased with age, from 1·2% in cases aged 40-49 years to 9·1% in patients aged ≥70 years. In line with previous reports from various settings, the strongest independent risk factors of deaths were older age, male sex, and the presence of one or more chronic comorbidities. Important novel findings were that sub-district-level socio-demographic factors, especially high population density and poverty, and health care factors, especially low COVID-19 vaccine coverage, further increased the risk of COVID-19-related death in metropolitan Jakarta.

A previous US study conducted in the early phase of the pandemic showed a significant association between household crowding and COVID-19 outcomes^18^; counties with the highest household crowding had a nearly 2-fold higher COVID-19 mortality rate than counties with the lowest crowding. Concordant with that study, we found that residents in sub-districts with the highest population density had a 34% higher risk of death than those residing in sub-districts with the lowest density. This finding typically relates to crowded urban communities who have the lowest standards of sanitation and waste management, and housing along flood-prone riverbanks. These sub-districts are also known to have relatively higher prevalence of non-communicable diseases such as hypertension and diabetes, and poverty-related infectious disease such as tuberculosis (Supplementary Table 5), which are well-established risk factors for worse COVID-19-associated clinical outcomes^28^. Reducing mortality in these areas may require comprehensive interventions such as improving diagnosis and case management of those known non-communicable and infectious diseases, as well as ensuring high COVID-19 vaccine coverage, and a sustainable social security network that may reduce vulnerability of these communities.

Socio-economic status including poverty has been associated with COVID-19 mortality in previous studies from South America^15–17^. In Chile, living in municipalities with lower socio-economic status was associated with increased risk of COVID-19-related mortality among general population^15^. In Brazil, living in a region with lower socio-economic status was associated with higher mortality risk among children hospitalised with COVID-19^16,17^. Similar to those studies, we found that the risk of death was 40% higher for resident of sub-districts with higher poverty (quarter 3) relative to those of lowest poverty. The interaction between poverty and population density also revealed that the risk of death was 30% higher for sub-districts with highest level of poverty and density compared to sub-districts with lowest poverty and density. Urban crowding and poverty impose very many disadvantages to health, here shown to include elevated risk of death as a consequence of SARS-CoV-2 infection.

The risk of COVID-19-related death in DKI Jakarta was 25% higher for resident of sub-districts with the lowest vaccine coverage (33-36%), compared to resident of sub-districts with the highest COVID-19 vaccine coverage (41-51%) as of August 31, 2021. This finding indicates that sub-districts with higher vaccine coverage can significantly reduce risk of mortality compared to those sub-districts with lower vaccine coverage. A previous study from Brazil reported that rapid scaling up of vaccination coverage among elderly Brazilians was associated with significant declines in relative mortality compared to younger individuals, in a setting where the gamma variant predominated^29^. Moreover, a recent modelling study estimated that the US states of Florida and Texas could have averted more than 95,000 hospital admissions and 22,000 deaths, if they had reached the vaccination coverage achieved by the top five states and continued at the same pace until August 31, 2021^30^. Those observations, corroborated in DKI Jakarta by the present study, highlight the health dividend of reduced mortality with rapid vaccination roll outs targeting the most vulnerable in reducing COVID-19-related deaths. As per November 23, 2021, two-dose COVID-19 vaccination coverage was 99·4% (8,887,770/8,941,211 targeted population) in DKI Jakarta, and 43·2% (89,892,161/208,265,720 targeted population) in Indonesia.

Consistent with evidence from previous studies across various settings, our findings affirm that older age, male sex and presence of underlying comorbidities were associated with higher risk of COVID-19-related mortality in the general population of DKI Jakarta, Indonesia^11–13,15,17^. The finding that children aged under 5 years old were 60% more likely to die during the first wave but not during the second, especially for those living in vulnerable districts, indicates that the youngest populations have been suffering most from gaps in access to health services, clinical management, and community support during the early epidemic response. Similar findings were reported from Brazil, where the risk of in-hospital death was 1·4-fold higher for children aged <2 years compared to those aged 2-11 years, those with comorbidities and living in areas with lower socio-economics status^16^. Hypertension and diabetes have each been associated with elevated risk of COVID-19 death in this setting^13^.

This study had some limitations. The retrospective design and reliance on routine surveillance data meant that, for some key baseline variables, data were incomplete or uniformly unavailable (e.g., type of comorbidities and disease severity classification). As in many other settings, the individual-level socio-demographics data were not recorded in the current Indonesia’s national database^22^. Comorbidities were often self-reported or could be under-diagnosed, potentially resulting in underreporting and hence underestimation of effect sizes. Details on supportive care and treatment received were also not available for this analysis. There are several other relevant socio-demographics variables such as human development index and public health development index that may represent population and health system vulnerability^24^ but were only available at the district level, and were not included in our analysis. However, our analysis included all available key variables that compose those indicators (prevalence of infectious and non-communicable diseases, health care workers-population ratio, universal child immunization, and all-cause mortality among under 5 years old population), therefore enhancing credibility of our findings.

In conclusion, individual-level risk factors associated with COVID-19 mortality in DKI Jakarta, Indonesia are broadly similar to those in more developed settings, dominated by advanced age and comorbidities. At the community-level, our analysis suggested that COVID-19 disproportionately affected people living in areas of high population density, poverty, and lower vaccination coverage. These findings indicate that vulnerability to death associated with COVID-19 includes not only the elderly and comorbid, but also the urban poor. This finding may inform decisions on health resource allocation against COVID-19 delivering the greatest possible health dividends by prioritising interventions, including vaccination, fort the most vulnerable communities. Future nationwide studies assessing individual, community, and health care capacity vulnerability associated with COVID-19-related mortality are needed to better understand the COVID-19 impact and to better tailor interventions to prioritise the most vulnerable communities.

## Data Availability

All data produced in the present study are available upon reasonable request to the authors

## Contributors

HS designed the study, did the analysis, and had full access to all of the data in the study and take responsibility for the integrity of the data and the accuracy of the data analysis. NS, VA, W, and DO did data collection and verification. KDL did the data cleaning. HS, AHS, RLH, JKB, and IRFE, contributed to the analysis and drafted the paper. All authors critically revised the manuscript for important intellectual content and all authors gave final approval for the version to be published.

## Data sharing statement

After publication, the datasets used for this study will be made available to others on reasonable requests to the corresponding author, including a detailed research proposal, study objectives and statistical analysis plan. Deidentified participant data will be provided after written approval from the corresponding author and the DKI Jakarta Health Office.

## Acknowledgments

This work was funded by the Wellcome (UK) Africa Asia Programme Vietnam (106680/Z/14/Z). We acknowledge all health care workers involved in the care for the COVID-19 patients, as well as those involved in the field data collection. Data cleaning was supported by funding from the Open Society Foundations.

## Supplementary data

**Table S1:** Age-specific mortality with confidence limits in COVID-19 cases in Jakarta, Indonesia, March 2, 2020 to August 31, 2021.

|  | N total | N deceased | % deceased | 95% CI |
| --- | --- | --- | --- | --- |
| Age group (years) |  |  |  |  |
| 0-4 | 21,793 | 47 | 0.22 | 0.16-0.29 |
| 5-9 | 23,070 | 16 | 0.07 | 0.04-0.11 |
| 10-19 | 66,514 | 72 | 0.11 | 0.09-0.14 |
| 20-29 | 149,267 | 311 | 0.21 | 0.19-0.23 |
| 30-39 | 146,900 | 692 | 0.47 | 0.44-0.51 |
| 40-49 | 121,454 | 1,455 | 1.20 | 1.14-1.26 |
| 50-59 | 98,934 | 3,115 | 3.15 | 3.04-3.26 |
| 60-69 | 52,776 | 2,827 | 5.36 | 5.17-5.55 |
| 70-79 | 24,261 | 2,261 | 9.13 | 8.78-9.50 |

**Table S2:** Sensitivity analysis of individual and community-level risk factors associated with COVID-19 mortality in Jakarta, Indonesia, March 2, 2020 to August 31, 2021. Table S2 presents the final multi-level multivariable logistic regression with multiple imputations for missing comorbidities. Sub-district was treated as the random effect variable. First wave: March 2020 to April 2021, Second wave: May 2021 to August 2021. Numeric values were categorised into quartiles (Q), i.e., below 25^th^ percentile (Lowest), 25^th^–50^th^ percentile (Q2), 50^th^–75^th^ percentile (Q3) and above 75^th^ percentile (Highest) for each sub-district level variable.

|  | Adjusted OR (95% CI) | p value |
| --- | --- | --- |
| **Individual factors** |  |  |
| Age group, years |  |  |
| 0-4 | 0.71 (0.44-1.14) | 0.154 |
| 5-9 | 0.30 (0.15-0.59) | <0.0001 |
| 10-19 | 0.50 (0.36-0.71) | <0.0001 |
| 20-29 | 1 (ref) | .. |
| 30-39 | 1.97 (1.64-2.36) | <0.0001 |
| 40-49 | 4.41 (3.73-5.22) | <0.0001 |
| 50-59 | 11.75 (10.03-13.76) | <0.0001 |
| 60-69 | 17.22 (14.65-20.25) | <0.0001 |
| ≥70 | 30.53 (25.91-35.99) | <0.0001 |
| Sex |  |  |
| Female | 1 (ref) | .. |
| Male | 1.29 (1.24-1.34) | <0.0001 |
| Comorbidities |  |  |
| No | 1 (ref) | .. |
| Yes | 4.25 (3.81-4.75) | <0.0001 |
| Period of time |  |  |
| First wave | 0.87 (0.69-1.09) | 0.215 |
| Second wave | 1 (ref) | .. |
| **Community (sub-district)-level factors** |  |  |
| Population density |  |  |
| Lowest | 1 (ref) | .. |
| Q2 | 1.18 (1.03-1.35) | 0.017 |
| Q3 | 1.22 (1.06-1.40) | 0.005 |
| Highest | 1.32 (1.14-1.54) | <0.0001 |
| Poverty level |  |  |
| Lowest | 1 (ref) | .. |
| Q2 | 1.39 (1.15-1.67) | 0.001 |
| Q3 | 1.33 (1.16-1.52) | <0.0001 |
| Highest | 1.16 (0.97-1.40) | 0.109 |
| Vaccine coverage |  |  |
| Lowest | 1.23 (1.13-1.35) | <0.0001 |
| Q2 | 1.18 (1.09-1.28) | <0.0001 |
| Q3 | 1.03 (0.90-1.18) | 0.059 |
| Highest | 1 (ref) | .. |

**Table S3:** Multivariable analysis of effect of age on mortality associated with COVID-19 mortality in Jakarta, Indonesia, March 2, 2020 to August 31, 2021. Table S3 presents the time-specific effect of age on mortality, after controlling for sex, comorbidities, population density, poverty, and vaccine coverage, and correlation at sub-district level (random effect variable). OR=odds ratio. First wave=March 2020 to April 2021. Second wave=May 2021 to August 2021.

|  | First wave |  | Second wave |  |
| --- | --- | --- | --- | --- |
|  | Adjusted OR (95% CI) | p value | Adjusted OR (95% CI) | p value |
| Age group, years |  |  |  |  |
| **0-4** | **1.56 (1.04-2.35)** | **0.033** | **0.72 (0.45-1.16)** | **0.183** |
| 5-9 | 0.41 (0.19-0.88) | 0.022 | 0.31 (0.16-0.61) | 0.001 |
| 10-19 | 0.53 (0.35-0.80) | 0.003 | 0.52 (0.37-0.72) | <0.0001 |
| 20-29 | 1 (ref) | .. | 1 (ref) | .. |
| 30-39 | 2.48 (2.03-3.04) | <0.0001 | 1.97 (1.64-2.36) | <0.0001 |
| 40-49 | 6.87 (5.70-8.29) | <0.0001 | 4.50 (3.80-5.32) | <0.0001 |
| 50-59 | 17.72 (14.80-21.22) | <0.0001 | 12.33 (10.52-14.44) | <0.0001 |
| 60-69 | 32.81 (27.39-39.29) | <0.0001 | 18.37 (15.64-21.58) | <0.0001 |
| ≥70 | 58.99 (49.11-70.86) | <0.0001 | 33.45 (28.41-39.38) | <0.0001 |

**Table S4:** Effect of poverty level on COVID-19 mortality based on population density. Table S4 presents the effect of poverty on mortality based on population density, after controlling for sex, comorbidities, time period, vaccine coverage, and correlation at sub-district level (random effect variable). OR=odds ratio. First wave=March 2020 to April 2021. Second wave=May 2021 to August 2021. First wave: March 2020 to April 2021, Second wave: May 2021 to August 2021.

|  | Adjusted OR (95% CI) | p value |
| --- | --- | --- |
| Lowest density |  |  |
| Lowest poverty | 1 (ref) | .. |
| Q2 | 1.43 (1.17-1.75) | <0.0001 |
| Q3 | 1.35 (1.34-0.77) | <0.0001 |
| Highest poverty | 1.18 (0.97-1.43) | 0.093 |
| Q2 |  |  |
| Lowest poverty | 1.20 (1.04-1.39) | 0.012 |
| Q2 | 1.12 (0.94-1.34) | 0.192 |
| Q3 | 1.34 (1.16-1.55) | <0.0001 |
| **Highest poverty** | **1.25 (1.08-1.45)** | **0.003** |
| Q3 |  |  |
| Lowest poverty | 1.23 (1.06-1.42) | 0.006 |
| Q2 | 1.37 (1.18-1.58) | <0.0001 |
| Q3 | NA | NA |
| **Highest poverty** | **1.23 (1.05-1.45)** | **0.011** |
| Highest density |  |  |
| Lowest poverty | 1.34 (1.14-1.58) | <0.0001 |
| Q2 | 1.42 (1.15-1.75) | 0.001 |
| Q3 | 1.24 (1.06-1.45) | 0.008 |
| **Highest poverty** | **1.27 (1.10-1.47)** | **0.001** |

**Table S5:** Association between population density with poverty, prevalence of hypertension, diabetes, and tuberculosis. Association between population density and other covariates. Numeric values were categorised into quartiles (Q), i.e., below 25^th^ percentile (Lowest), 25^th^–50^th^ percentile (Q2), 50^th^–75^th^ percentile (Q3) and above 75^th^ percentile (Highest) for each sub-district level variable.

|  | Population density |  |  |  |  |
| --- | --- | --- | --- | --- | --- |
|  | Lowest | Q2 | Q3 | Highest | p value |
| Poverty |  |  |  |  |  |
| Lowest | 35.18% | 28.24% | 27.84% | 22.53% | <0.0001 |
| Q2 | 11.05% | 13.14% | 49.16% | 11.14% |  |
| Q3 | 44.38% | 28.27% | 0.00% | 31.02% |  |
| Highest | 9.38% | 30.34% | 23.00% | **35.32%** |  |
| Hypertension prevalence |  |  |  |  |  |
| Lowest | 27.48% | 27.57% | 25.19% | 17.21% | <0.0001 |
| Q2 | 27.30% | 34.71% | 29.29% | 22.90% |  |
| Q3 | 10.88% | 22.63% | 18.42% | 30.62% |  |
| Highest | 34.34% | 15.09% | 27.10% | **29.27%** |  |
| Diabetes prevalence |  |  |  |  |  |
| Lowest | 14.23% | 39.26% | 34.31% | 10.24% | <0.0001 |
| Q2 | 34.75% | 32.97% | 0.00% | 26.33% |  |
| Q3 | 31.75% | 5.38% | 29.75% | 38.09% |  |
| Highest | 19.27% | 22.39% | 35.94% | **25.33%** |  |
| Tb prevalence |  |  |  |  |  |
| Lowest | 60.29% | 0.00% | 19.56% | 20.15% | <0.0001 |
| Q2 | 31.09% | 27.13% | 21.23% | 20.54% |  |
| Q3 | 5.77% | 28.43% | 40.62% | 25.19% |  |
| Highest | 9.12% | 50.77% | 12.65% | **27.45%** |  |

**Table S6:** Association between poverty with prevalence of hypertension, diabetes, and tuberculosis. Association between poverty and other covariates. Numeric values were categorised into quartiles (Q), i.e., below 25^th^ percentile (Lowest), 25^th^–50^th^ percentile (Q2), 50^th^–75^th^ percentile (Q3) and above 75^th^ percentile (Highest) for each sub-district level variable.

|  | Poverty |  |  |  |  |
| --- | --- | --- | --- | --- | --- |
|  | Lowest | Q2 | Q3 | Highest | p value |
| Hypertension prevalence |  |  |  |  |  |
| Lowest | 16.94% | 46.95% | 27.97% | 10.15% | <0.0001 |
| Q2 | 29.16% | 41.99% | 12.34% | 33.75% |  |
| Q3 | 42.88% | 0.00% | 25.97% | 8.09% |  |
| Highest | 11.03% | 11.06% | 33.71% | **48.01%** |  |
| Diabetes prevalence |  |  |  |  |  |
| Lowest | 13.43% | 54.05% | 27.97% | 13.43% | <0.0001 |
| Q2 | 28.46% | 27.44% | 20.38% | 14.55% |  |
| Q3 | 40.26% | 0.00% | 17.94% | 37.25% |  |
| Highest | 17.85% | 18.50% | 33.71% | **34.77%** |  |
| Tb prevalence |  |  |  |  |  |
| Lowest | 37.44% | 0.00% | 45.83% | 15.67% | <0.0001 |
| Q2 | 38.55% | 11.32% | 19.29% | 33.83% |  |
| Q3 | 11.03% | 53.05% | 7.68% | 30.56% |  |
| Highest | 12.99% | 35.63% | 27.21% | **19.94%** |  |

**Figure S1.**
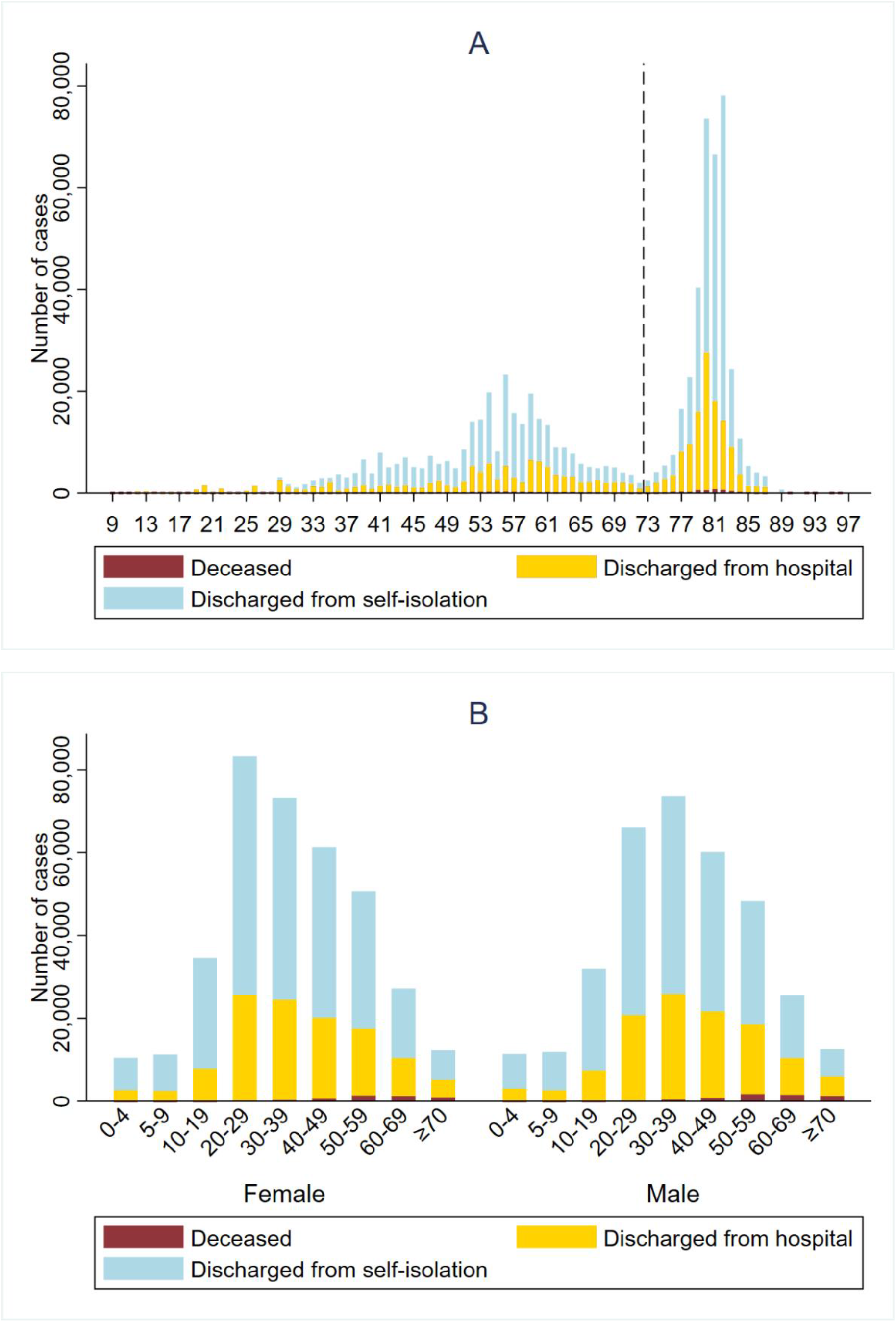
Outcomes over time (A), over age and by gender (B). Dashed black line represents the cut-off between the first and the second wave.

